# Pod-based e-cigarette use among college-aged adults in the United States: A survey on the perception of health effects, sociodemographic correlates, and interplay with other tobacco products

**DOI:** 10.1101/2022.10.22.22281396

**Authors:** Olufunmilayo H. Obisesan, S.M. Iftekhar Uddin, Ellen Boakye, Albert D. Osei, Mohammadhassan Mirbolouk, Olusola A. Orimoloye, Omar Dzaye, Omar El Shahawy, Andrew Stokes, Andrew P. DeFilippis, Emelia J. Benjamin, Michael J. Blaha

## Abstract

**Introduction:** E-cigarette use among youth and young adults remains of public health concern. Pod-based e-cigarettes, including JUUL, significantly changed the e-cigarette landscape in the United States (US). Using an online survey, we aimed to explore the socio-behavioral correlates, predisposing factors, and addictive behaviors among young adult pod-mod users within a University in Maryland, USA.

**Methods:** 112 eligible college students aged 18-24 years recruited from a University in Maryland who reported using pod-mods were included in this study. Participants were categorized into current/non-current users based on past-30-day use. Descriptive statistics were used to analyze participants’ responses.

**Results:** Of the 112 participants with mean age 20.5±1.2 years, 56.3% were female, 48.2% White, and 40.2% reported past-30-day (current) use of pod-mods. The mean age of first experimentation with pod-mods was 17.8±1.4 years, while the mean age of regular use was 18.5±1.4 years, with the majority (67.9%) citing social influence as the reason for initiation. Of current users, 62.2% owned their own devices, and 82.2% predominantly used JUUL and menthol flavor (37.8%). A significant proportion of current users (73.3%) reported buying pods in person, 45.5% of whom were below the age of 21. Among all participants, 67% had had a past serious quit attempt. Among them, 89.3% neither used nicotine replacement therapy nor prescription medications. Finally, current use (adjusted odds ratio (aOR):4.52; 95%CI:1.76,11.64), JUUL use (aOR:2.56; 95%CI:1.08,6.03), and menthol flavor (aOR:6.52; 95%CI:1.38,30.89) were associated with reduced nicotine autonomy, a measure of addiction.

**Conclusion:** Our findings provide specific data to inform the development of public health interventions targeted at college youth, including the need for more robust cessation support for pod-mod users.

## Introduction

The study of electronic nicotine delivery systems (ENDS) use among young individuals is important for many reasons. First, ENDS use has been associated with many short-term health conditions affecting several organ systems, including the respiratory and cardiovascular systems, as well as mental health conditions such as depression.^1–4^ Secondly, ENDS contain widely varying levels of nicotine and have been associated with subsequent cigarette smoking and other risky health behaviors including use of non-prescription drugs, marijuana, and heavy alcohol.^5–8^ Most importantly, although promoted as a tobacco-cessation tool, population surveillance has shown significant use among young tobacco naïve individuals, many of whom report frequent use.^9–11^

Of particular concern are the rapidly changing design and the innovation of new ENDS products that continue to be introduced into the market. Pod-based e-cigarettes, also known as pod-mods, were introduced onto the market in 2015 and quickly became the dominant product in the e-cigarette space due to their sleekness, ease of concealment, and versatility.^7^ JUUL, a prominent pod-mod e-cigarette, fueled the youth e-cigarette epidemic due to youth-targeted advertisement, and until recently, was the most widely used e-cigarette brand among youth and young adults in the US. ^12,13^

To address the widespread uptake of ENDS among youth, the Food and Drug Administration (FDA) and the Federal government implemented policies such as increasing the age of sale of tobacco products from 18 to 21 years in 2019 and the ban on flavors in 2020.^14,15^ Additionally, more recently, the FDA has denied authorization to market JUUL products in the US.^16^ Population surveys show that in 2020, 5.1% of US adults reported using ENDS, a prevalence largely driven by young adults aged 18-24 years, especially students.^10^ Among college students, lifetime e-cigarette use has been reported to be as high as 36.0% - 39.8%.^17^ This age demographic is crucial because they represent the transitional period between adolescence and adulthood, a period defined by changing social roles and associated with experimentation and risk-taking behavior.^17,18^ Additionally, individuals in this age range are the youngest legal targets of advertisements by the tobacco industry and are highly responsive to social influence.^7,19^ It is, therefore, crucial to delineate the correlates of pod-mod use in this highly susceptible group of young adults.

To understand the sociodemographic correlates and the predisposing factors associated with pod-based e-cigarette use among young college-aged adults in the United States, and to evaluate typical use patterns and addictive behaviors among this demographic, we utilized an online survey tool administered to confirmed students aged 18-24 years in a University in Maryland, USA, who reported use of pod-mods. We also sought to understand the perceptions of harm associated with ENDS use juxtaposed with awareness about public health messages on tobacco use.

## Methods

### Study Participants/ Eligibility criteria

Participants were confirmed students from a large urban University in Maryland, USA, within the age group of 18-24 years who reported use of pod-based e-cigarettes. Participants were recruited from September 2020 to March 2021 via regular announcements on the University’s media hub announcement page. Of the 604 individuals who indicated an interest in the study, 471 could not verify their student status and, thus, did not meet the eligibility criteria. The remaining 133 individuals received unique identifier numbers and a link to the survey, of whom 117 completed the survey. Among them, 5 individuals had inconsistent entries and were eliminated from the analyses. Therefore, 112 valid survey responses were analyzed.

### Study Procedures

An online survey was designed on the University’s REDCap platform, a secure web application. The survey was designed to be easily understandable, with a Flesch-Kincaid grade level score of 7. Questions were developed based on the principles of the Integrated Behavioral Model.^20,21^ When possible, questions were modeled after questions in the PhenX toolkit (an online catalog of scientifically validated measures related to a wide range of research domains, including tobacco regulatory research).^22^ Others were modeled after prior published work (Supplement 6).^23–27^ In total,75 questions were asked, grouped under the following domains: demographic information, general tobacco and other nicotine products use history, patterns of pod-mod use, perceptions of potential harms associated with vaping, and awareness of public health messages on vaping.

Approval for the study was obtained from the Johns Hopkins University’s Institutional Review Board. To protect the identity of participants, their communication with the study team was via a designated member of the team with no access to the data. Participants’ student status was verified, and they were required to take an honor pledge that they met other eligibility criteria, following which they were given unique identification numbers (UIN). With these UINs, participants could access the survey where they were presented with informed consent for the study. After consenting to the study procedures, they could then take the survey. Upon completion, they received a $45 incentive and were asked for referrals of other students using pod-mod e-cigarettes who would be interested in taking the survey. Data from the study were stored and analyzed within a HIPAA compliant environment on the University’s SafeDesktop.

### Measures

#### Demographic characteristics

Participants were asked about their age, sex, race/ethnicity, and household income.

#### Pod-mod use patterns

An introduction about pod-mods and examples of brands were provided, together with a link to pictures. Participants were categorized based on their response to questions about past 30-day pod-mod use. Participants who reported past-30-day use of pod-mods were categorized as *current users*, and those who reported no use of pod-mods in the past 30-days were *non-current users*. Preferred flavors and brands were also assessed, and participants were asked about the number of pods they typically finished in a month.

A brief introduction about other tobacco products was also provided, with a link to pictures. Participants were then asked about lifetime use and frequency of use of other nicotine products including cigarettes, cigars, cigarillos, pipes, hookah, smokeless tobacco (snus pouches, loose snus, moist snuff, dip, spit or chewing tobacco), and dissolvable tobacco. They were also asked about marijuana use.

#### Predisposing factors and reasons for use

To assess predisposing factors for initial and continued use, participants were asked questions about ages of initial and regular use. They were also asked about ownership of pod-mods and purchase patterns. To assess perceived social influence and acceptability of pod-mod use, the number of family/close friends who used pod-mods and their opinions about pod-mod use were asked. Questions were also asked to assess exposure to pod-mod marketing.

To assess reasons for initiating pod-mod use, participants were asked “why did you start using pod-mods”, several options were provided some of which include: “I was curious”, “Friends/Family use or gave me one to try”, “They are less harmful to me compared to smoking regular cigarettes”, “They helped me quit/reduce smoking” and multiple answers were allowed. Participants were also asked about where they first tried pod-mods.

#### Addictive behaviors, Autonomy, and Quit Attempts

To assess addictive behaviors, participants were asked questions including: “how soon after waking up do/did you take your first puff of a pod-mod” and “Have you ever felt like you were addicted to using pod-mods”. Given that diminished autonomy is peculiar to all forms of drug or behavior dependence,^28^ we adapted the hooked on nicotine checklist (HONC), a reliable and valid measure of diminished autonomy over tobacco, to assess participants’ addictive behaviors.^24,29^ Participants were categorized into 2 groups: those with full autonomy (HONC score of 0) and those with reduced autonomy (HONC score≥1).

To assess characteristics associated with quit attempts, number of prior quit attempts, reasons for quitting, and symptoms experienced when they attempted to quit were explored.

#### Perceptions of potential harms

Perceptions of potential harms were assessed with questions including: “what in your view are the main harms, if any, of pod-mod use”. Multiple answers were allowed and some of the options provided were “there are no harms” and “there has not been enough research done to understand all the possible harms”.

#### Exposure to public health messages on vaping

Participants were asked if they had seen any anti-smoking or anti-tobacco ads on TV or social media. Their opinions about the ads were assessed using a Likert scale from “strongly agree” to “strongly disagree”.

### Data Analysis

Sociodemographic characteristics of participants were analyzed using descriptive analyses and data were presented using means/medians and proportions. Using descriptive analyses, current and non-current users were further characterized per the domains listed earlier.

The association between nicotine autonomy (HONC) and pod-mod brands, flavors, and the use of other nicotine products was assessed using descriptive analyses and logistic regression models adjusted for age and sex.

All analyses were conducted using Stata software version 15.1 (Stata Corp, College Station, TX), with statistical significance set using a two-sided p-value< 0.05.

## Results

There were 112 eligible participants, of whom 59.8% were non-current users and 40.2% were current users. Participants had a mean age of 20.5years (SD:1.2years) and were predominantly female (56.3%), White (48.2%), non-Hispanic (83%), with household income above $100,000 (61.6%). Compared to non-current users, a larger proportion of current users were male, White, and Hispanic. Other demographic characteristics were similar between both groups (Table 1).

**Table 1:** Demographic characteristics of participants

|  | N (%) |  |  |
| --- | --- | --- | --- |
| Characteristic | Total<br>(N=112) | Current user<br>(N=45) | Non-current user<br>(N=67) |
| Age (mean (SD)) | 20.5(1.2) | 20.6(1.2) | 20.4(1.2) |
| Sex |  |  |  |
| Female | 63(56.3) | 21(46.7) | 42(62.7) |
| Male | 49(43.8) | 24(53.3) | 25(37.3) |
| Race |  |  |  |
| Asian | 42(37.5) | 14(31.1) | 28(41.8) |
| Black or African American | 8(7.1) | 3(6.7) | 5(7.5) |
| White | 54(48.2) | 24(53.3) | 30(44.8) |
| Multiracial | 4(3.6) | 2(4.4) | 2(3.0) |
| Other | 4(3.6) | 2(4.4) | 2(3.0) |
| Ethnicity |  |  |  |
| Hispanic | 19(17.0) | 12(26.7) | 7(10.5) |
| Non-Hispanic | 93(83.0) | 33(73.3) | 60(89.6) |
| Household income |  |  |  |
| <\$35,000 | 5(4.5) | 2(4.4) | 3(4.5) |
| \$35,000 - \$50,000 | 6(5.4) | 1(2.2) | 5(7.5) |
| \$50,000 - \$75,000 | 11(9.8) | 9(20.0) | 2(3.0) |
| \$75,000 - \$100,000 | 13(11.6) | 5(11.1) | 8(11.9) |
| >\$100,000 | 69(61.6) | 26(57.8) | 43(64.2) |
| Unsure | 8(7.1) | 2(4.4) | 6(9.0) |

### Pod-Mod use characteristics

Current users had higher average monthly pod consumption rates compared to non-current users when they were using pod-mods. Current users were also more likely to live with people who used pod-mods and have more friends who also used pod-mods. A larger proportion of current users also reported having tried multiple tobacco products - 82.2% had tried combustible cigarettes, 42.2% tried cigarillos, and 51.1% had tried hookah compared to 50.8%, 22.4%, and 29.9% of non-current users for each product, respectively. Only 2.2% of participants reported concurrent daily use of combustible cigarettes (Table 2). Overall, most participants reported that their close contacts had ambivalent (36.6%) or negative (48.2%) opinions about pod-mods. Their perception of the general public’s opinion of pod-mods followed a similar pattern (Table 2).

**Table 2:** Pod mod use characteristics

|  | N (%) |  |  |  |
| --- | --- | --- | --- | --- |
|  | Total<br>(N=112) | Current users<br>(N=45) | Non-current<br>users<br>(N=67) | p-value |
| Age first tried pod-mod (mean(SD)) | 17.8(1.4) | 17.8(1.3) | 17.8(1.4) | 0.80 |
| Age commenced regular use (mean(SD)) | 18.5(1.4) | 18.8(1.3) | 18.3(1.4) | 0.10 |
| First pod flavor used |  |  |  | 0.05 |
| Virginia tobacco | 1(0.9) | 0(0.0) | 1(1.5) |  |
| Mint | 41(36.6) | 21(46.7) | 20(29.9) |  |
| Mango | 28(25) | 9(20.0) | 19(28.4) |  |
| Crème | 5(4.5) | 1(2.2) | 4(6.0) |  |
| Cucumber | 3(2.7) | 3(6.7) | 0(0.0) |  |
| Classic tobacco | 2(1.8) | 2(4.4) | 0(0.0) |  |
| Menthol | 8(7.1) | 3(6.7) | 5(7.5) |  |
| Fruit | 17(15.2) | 6(13.3) | 11(16.4) |  |
| Other | 5(4.5) | 0(0.0) | 5(7.5) |  |
| None | 2(1.8) | 0(0.0) | 2(3.0) |  |
| Average monthly pod consumption |  |  |  | 0.01 |
| Less than 1 pod | 54(48) | 15(33.3) | 39(58.2) |  |
| 1 to 2 pods | 14(12.5) | 6(13.3) | 8(11.9) |  |
| 3 to 4 pods | 17(15.2) | 6(13.3) | 11(16.2) |  |
| 5 to 12 pods | 14(12.5) | 9(20.0) | 5(7.5) |  |
| 13 to 19 pods | 7(6.3) | 3(6.7) | 4(6.0) |  |
| 20 or more | 6(5.4) | 6(13.3) | 0(0.0) |  |
| Living with people who use pod-mods | 37(33.0) | 24(53.3) | 13(19.4) | 0.00 |
| Of 5 closest friends, number who use pod<br>mods |  |  |  | 0.01 |
| 0 | 16(14.3) | 2(4.4) | 14(20.9) |  |
| 1 | 34(30.4) | 10(22.2) | 24(35.8) |  |
| 2 | 29(25.9) | 12(26.7) | 17(25.4) |  |
| 3 | 19(17.0) | 11(24.4) | 8(11.9) |  |
| 4 | 9(8.0) | 7(15.6) | 2(3.0) |  |
| 5 | 5(4.5) | 3(6.7) | 2(3.0) |  |
| Opinion of close contacts about pod-mods |  |  |  | 0.59 |
| Positive | 17(15.2) | 6(13.3) | 11(16.4) |  |
| Neither positive nor negative | 41(36.6) | 19(42.2) | 22(32.8) |  |
| Negative | 54(48.2) | 20(44.4) | 34(50.8) |  |
| Perception of general public's opinion about using pod-mods |  |  |  | 0.19 |
| Positive | 12(10.7) | 2(4.4) | 10(14.9) |  |
| Neither positive nor negative | 46(41.1) | 21(46.7) | 25(37.3) |  |
| Negative | 54(48.2) | 22(48.9) | 32(47.8) |  |
| Other tobacco product use |  |  |  |  |
| Combustible cigarette |  |  |  |  |
| Ever use | 71(63.4) | 37(82.2) | 34(50.8) | 0.00 |
| Current occasional use | 18(16.1) | 10(22.2) | 8(11.9) | 0.15 |
| Current daily use | 1(2.2) | 1(2.2) | 0(0.0) | 0.15 |
| Traditional cigar/Cigarillo/Blunt |  |  |  |  |
| Ever use (Cigar) | 44(39.3) | 21(46.7) | 23(34.3) | 0.19 |
| Ever use (Cigarillo) | 34(30.4) | 19(42.2) | 15(22.4) | 0.03 |
| Ever use (Blunts) | 67(59.8) | 32(71.1) | 35(52.2) | 0.05 |
| Current occasional use (Cigar/Cigarillo/Blunt) | 27(24.1) | 11(24.4) | 16(23.9) | 0.95 |
| Pipe |  |  |  |  |
| Ever use | 20(17.9) | 11(24.4) | 9(13.4) | 0.14 |
| Current occasional use | 2(1.8) | 1(2.2) | 1(1.5) | 0.69 |
| Current daily use | 1(0.9) | 0(0.0) | 1(1.5) | 0.69 |
| Hookah |  |  |  |  |
| Ever use | 43(38.4) | 23(51.1) | 20(29.9) | 0.02 |
| Current occasional use | 12(10.7) | 4(8.9) | 8(11.9) | 0.61 |
| Smokeless tobacco<sup>a</sup> |  |  |  |  |
| Ever use (snus pouches) | 6(5.4) | 2(4.4) | 4(6.0) | 0.73 |
| Ever use (loose snus/moist snuff) | 6(5.4) | 3(6.7) | 3(4.5) | 0.61 |
| Current daily use | 2(1.8) | 0(0.0) | 2(3.0) | 0.24 |

<sup>a</sup>Snus pouches, loose snus, moist snuff, dip, spit, or chewing tobacco

### Initiation of use

The mean age of first experimentation with pod-mods was 17.8years (SD:1.4 years), while the mean age of regular use was 18.5years (SD:1.4 years). Figure 1 shows the distribution of participants’ response to the question about their reasons for initiating pod-mod use. 67.9% reported initiation because “friends/family use or gave me one to try”, 62.2% reported being curious, and 25.9% tried them for the flavors. Majority reported first trying pod-mods at home or at a friend’s home (44.6%) or at a social gathering (party/nightclub/concert) (39.3%) (Figure 1).

**Figure 1:**
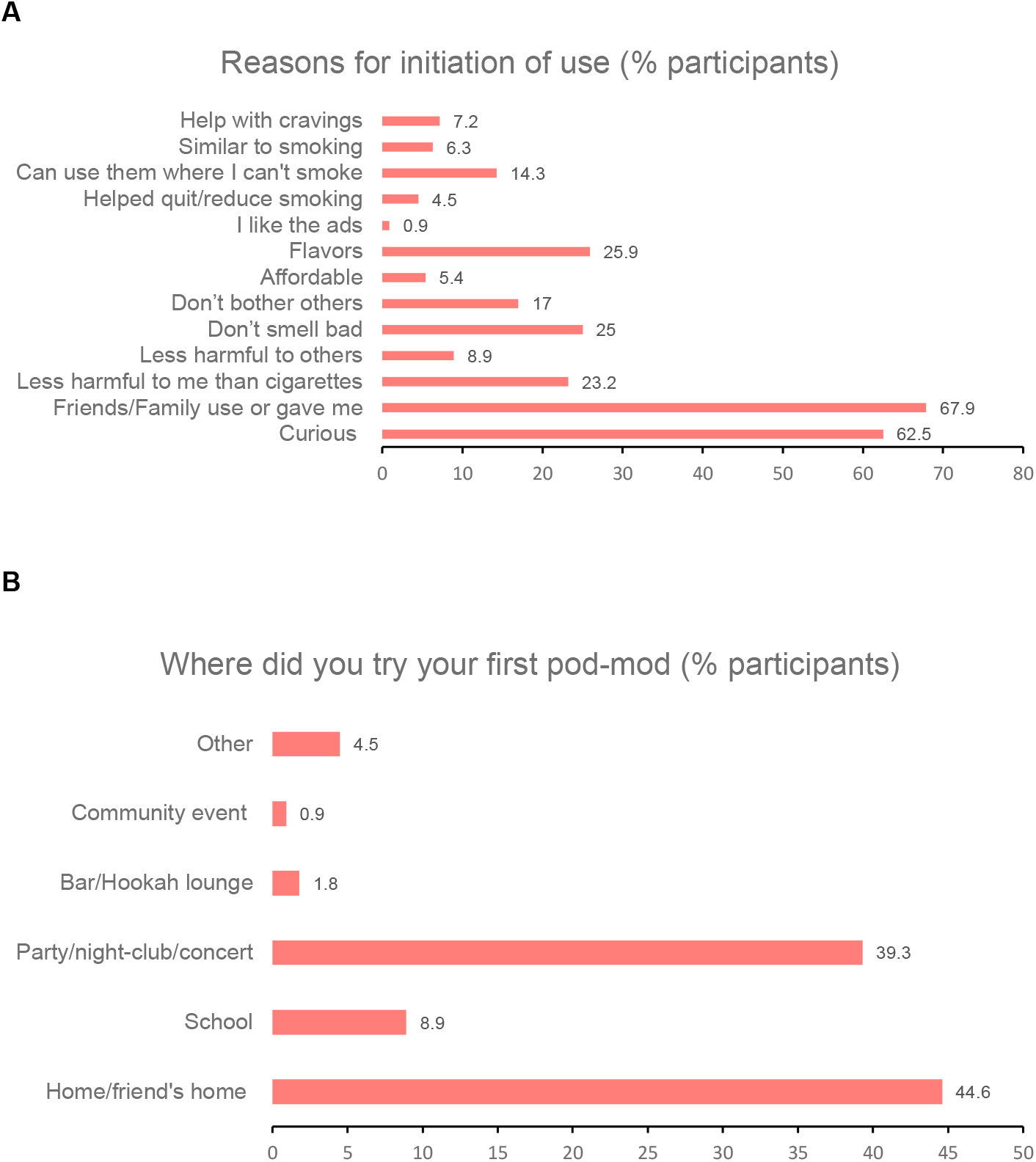
Initiation of use

### Characteristics of current users

Among current users, 82.2% reported using JUUL most frequently in the past month, menthol was the most frequently used flavor (37.8%), and 62.2% owned their own pod-mods. A total of 26.7% had been advised by health professionals to quit in the past year, and 68.8% rarely or never read the health warnings on pod-mod packaging (Table 3). The majority (73.3%) reported buying pods in person, 45.5% of whom were below the age of 21.

**Table 3:** Characteristics of current users

|  | N (%) |
| --- | --- |
| Characteristic | Current user, (N=45) |
| Most frequently used brand of pod e-cigarettes* |  |
| • JUUL | 37(82.2) |
| • Suorin | 2(4.4) |
| • Puff Bar | 3(6.7) |
| • Vuse | 1(2.2) |
| • Other | 2(4.4) |
| Pod flavor used most frequently* |  |
| • Virginia tobacco | 2(4.4) |
| • Mint | 9(20) |
| • Mango | 4(8.9) |
| • Menthol | 17(37.8) |
| • Fruit | 7(15.6) |
| • None | 4(8.9) |
| Own a pod-mod | 28(62.2) |
| Amount paid for pod-mods |  |
| • <\$10 | 2(4.4) |
| • \$10 - \$30 | 21(46.7) |
| • \$31 - \$40 | 6(13.3) |
| • \$41 - \$50 | 6(13.3) |
| • >\$50 | 2(4.4) |
| Method of purchase of pods |  |
| • In person | 33(73.3) |
| • From the internet | 2(4.4) |
| • Do not purchase my own | 10(22.2) |
| Health professional advised to quit using pod-mods in last 12 months | 12(26.7) |
| In the past 30 days, how often have you read the health warnings on pod-mod packages |  |
| • Never | 20(44.4) |
| • Rarely | 11(24.4) |
| • Sometimes | 12(26.7) |
| • Often | 2(4.4) |
| • Very often | 0(0.0) |

\*in the last 30 days

### Addictive behaviors and quit attempts

Some of the addictive behaviors reported by participants include having strong cravings for pod-mods (45.5%), feeling of being addicted (36.6%), having strong urges to use them during quit attempts (41.1%), feeling nervous/anxious during quit attempts (32.1%), and difficulty concentrating during quit attempts (23.2%). (Table 4) A larger proportion of current users reported addictive behaviors compared to non-current users, and 82.2% of current users had plans of quitting pod-mod use in the next 6 months compared to 59.7% of non-current users. Overall, 67% of participants had a past serious quit attempt (Table 4).

**Table 4:**
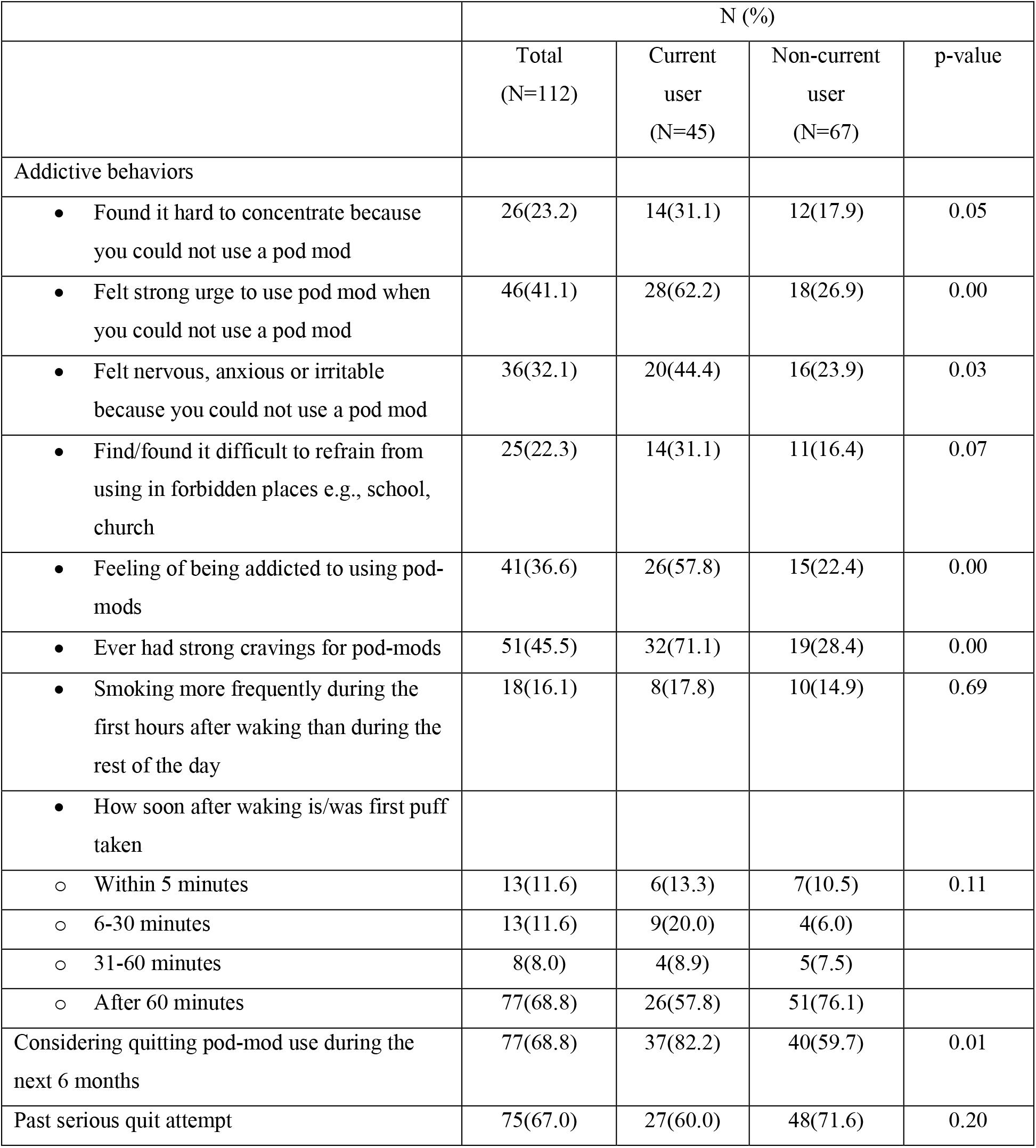
Addictive behaviors and quit attempts

|  | N (%) |  |  |  |
| --- | --- | --- | --- | --- |
|  | Total<br>(N=112) | Current<br>user<br>(N=45) | Non-current<br>user<br>(N=67) | p-value |
| Addictive behaviors |  |  |  |  |
| • Found it hard to concentrate because you could not use a pod mod | 26(23.2) | 14(31.1) | 12(17.9) | 0.05 |
| • Felt strong urge to use pod mod when you could not use a pod mod | 46(41.1) | 28(62.2) | 18(26.9) | 0.00 |
| • Felt nervous, anxious or irritable because you could not use a pod mod | 36(32.1) | 20(44.4) | 16(23.9) | 0.03 |
| • Find/found it difficult to refrain from using in forbidden places e.g., school, church | 25(22.3) | 14(31.1) | 11(16.4) | 0.07 |
| • Feeling of being addicted to using pod-mods | 41(36.6) | 26(57.8) | 15(22.4) | 0.00 |
| • Ever had strong cravings for pod-mods | 51(45.5) | 32(71.1) | 19(28.4) | 0.00 |
| • Smoking more frequently during the first hours after waking than during the rest of the day | 18(16.1) | 8(17.8) | 10(14.9) | 0.69 |
| • How soon after waking is/was first puff taken |  |  |  |  |
| ○ Within 5 minutes | 13(11.6) | 6(13.3) | 7(10.5) | 0.11 |
| ○ 6-30 minutes | 13(11.6) | 9(20.0) | 4(6.0) |  |
| ○ 31-60 minutes | 8(8.0) | 4(8.9) | 5(7.5) |  |
| ○ After 60 minutes | 77(68.8) | 26(57.8) | 51(76.1) |  |
| Considering quitting pod-mod use during the next 6 months | 77(68.8) | 37(82.2) | 40(59.7) | 0.01 |
| Past serious quit attempt | 75(67.0) | 27(60.0) | 48(71.6) | 0.20 |

Among participants with quit attempts, majority tried quitting because they were concerned about potential health risks (72%), did not feel like using them any longer (41.3%), or because of the expensive cost (42.7%). Majority of participants who had tried quitting (89.3%) neither used nicotine replacement therapy nor prescription medications such as bupropion during quit attempts (Table 5).

**Table 5:**
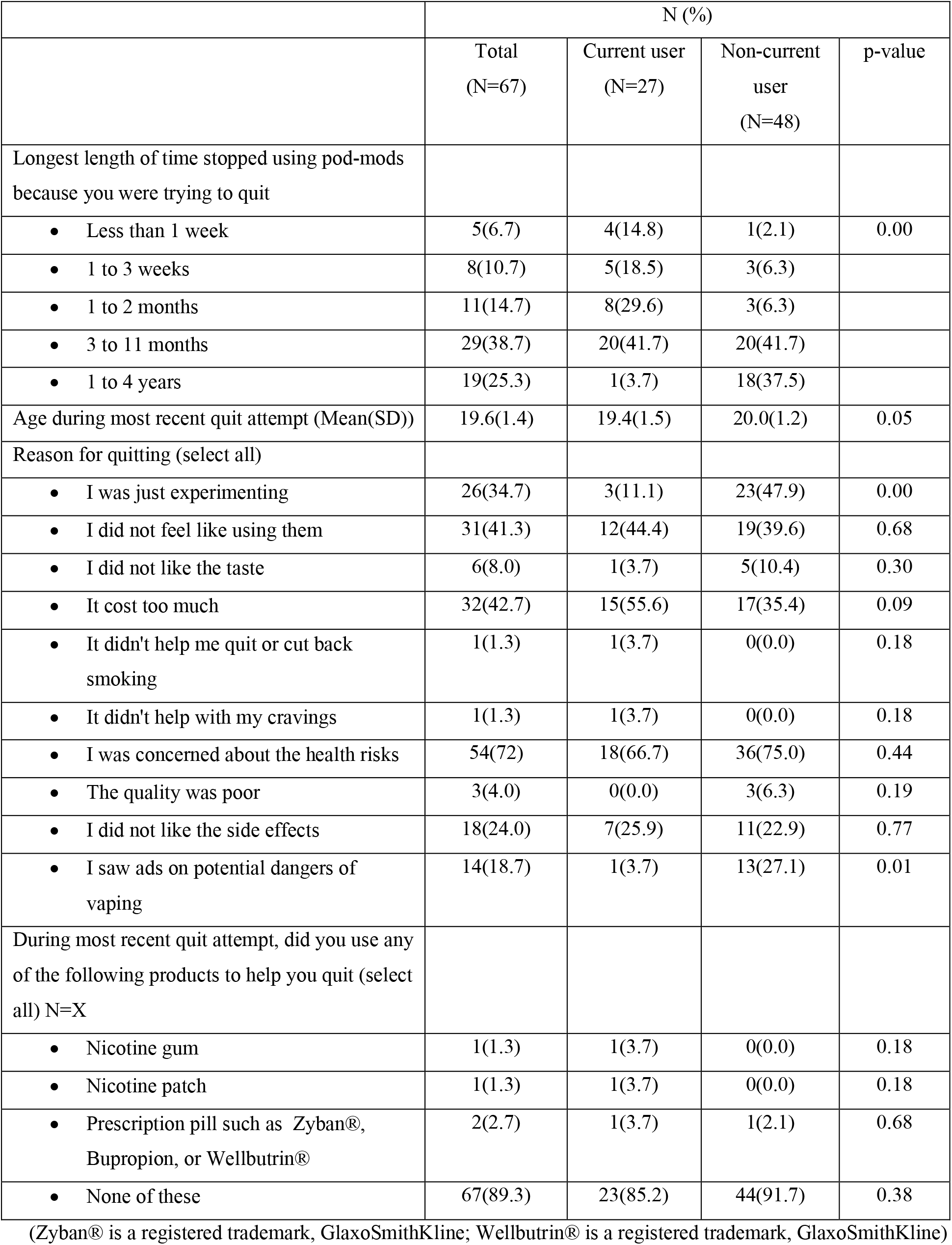
Characteristics associated with quit attempts

### Exposure to marketing/public health messages and perception of harm

Only a minority (17%) of participants reported seeing internet ads for pod-mods sometimes or most times, while 42.9% had never seen internet ads for them. However, 39.3% reported discussing pod-mods on their social networking accounts (Supplement Table 1). Although 74.1% of participants had seen anti-tobacco ads, only a minority (14.3%) had discussed the ad content with anyone. Approximately 35% thought the anti-tobacco ads were worth remembering; 39.3% agreed they were informative, and 27.7% thought they were convincing (Supplement Table 2).

Approximately 96% of participants were aware that pod-mods contain nicotine, 58% thought they were less harmful than cigarettes, and 47.3% thought they were less addictive than cigarettes. The most perceived harms associated with pod-mod use included: addiction (86.6%), respiratory problems (83.9%), harmful e-liquid constituents (75.9%), reinforcing of smoking habit (64.3%), and cancer (62.5%). Additionally, 74.1% of participants thought that not enough research had been done to understand all the harms associated with pod-mod use (Supplement Table 3).

### Characteristics of participants by nicotine autonomy status

Participants with reduced autonomy were more likely to be current users (61.3%), have JUUL as their most frequently used brand (62.9%), used menthol flavor most frequently (29%), and were more likely to have tried other tobacco products-cigarettes (77.4%), cigarillo (40.3%), blunt (70.9%) and hookah (51.6%) (Supplement Table 4).

In logistic regression analyses, after adjusting for age and sex, current users had 4.52 times (95%CI:1.76,11.64) higher odds of having reduced autonomy compared to non-current users. Participants who used JUUL most frequently had 2.56 times (95%CI:1.08,6.03) higher odds of having reduced autonomy compared to participants who had no preferred brand of pod-mods, and participants who used menthol flavor most frequently had 6.52 times (95%CI:1.38,30.89) higher odds of having reduced autonomy compared to participants who had no preferred flavor of pods (Supplement table 5).

## Discussion

In this multi-faceted survey study of college-aged pod-mod users, we observed that current users were more likely to own their own pod-mods and use JUUL and menthol flavor pods most frequently. They were also more likely to have tried multiple tobacco products, use more pods on average per month, live with people and have more friends who use pod mods, and report dependence/addictive behaviors compared to non-current users. A considerable proportion of current users aged <21 years reported buying their devices in-person. Overall, majority of participants initiated use due to curiosity and social influence, had well-informed perceptions of the potential harms associated with pod-mod use, and were considering quitting due to health risks. Additionally, majority of the participants reported past year quit attempts without the use of NRT or prescription medications.

While pod-mods may contain fewer toxicants than cigarettes and older generation e-cigarettes, they can deliver much higher doses of nicotine to the user via nicotine salts, and prior studies have shown an association between their use and higher levels of nicotine dependence.^30–32^ Although most prior studies have focused on adolescents and youth,^30–32^ our findings among the unique subgroup of college students show a similar pattern, as current users in our study were more likely to report addictive behaviors. Also, based on questions adapted from the HONC,^29^ current users in our study were more likely to have reduced autonomy. Nicotine has been shown to affect cognitive development, with resultant alterations in working memory, attention span and development of the reward processing center in the developing brain.^33,34^ Additionally, preclinical studies using rat models have shown that adolescents tolerate higher levels of nicotine compared to adults, and have blunted nicotine withdrawal symptoms, which may further increase the adolescent’s vulnerability to unchecked nicotine use and result in the “gateway” effect of transitioning to combustible cigarettes as they get older.^33^ Given that the brain continues to develop well into adulthood,^35^ the exposure of young adults to such high levels of nicotine as are present in many pod-mods may be viewed as concerning. Many current users also used multiple tobacco products, which could further reinforce addictive behaviors.

Reflecting its dominant share of the ENDS market space in 2019,^36^ JUUL was the most commonly used brand of e-cigarettes among participants in our study. This is consistent with other studies conducted among young adults in the population,^37–39^ lending further credence to assertions that the explosion of ENDS use among youth between 2017 and 2019 was partly fueled by the brand,^13^ whose influence is apparent regardless of education level. Similarly, menthol was the most frequently used flavor among participants in our study, also reflecting the ENDS market space, where following the federal ban of other flavors from the market, the menthol market share increased from 13% in 2019 to 46% in 2020.^40^

Of note, JUUL and menthol flavor pod-mod use were associated with reduced autonomy among participants, suggesting a higher addiction potential for both products. This is not unexpected as JUUL pods contain nicotine concentrations as high as 40mg/pod (equivalent to a pack of cigarettes),^41^ and menthol has been reported to increase nicotine dependence and hamper smoking cessation especially among youth, Black and Hispanic individuals.^42–44^

In our study, pod-mod initiation was largely mediated by peer and family influence and curiosity, and continued use was associated with living with people or having friends who also used pod mods, regardless of perceived opinions of others about the product. Our findings among college students are consistent with other studies among youth, highlighting the importance of social influence in this demographic.^39,45^ Additionally, more participants reported discussing pod-mods on their social media platforms, while only few participants reported seeing internet ads of the products. This is particularly intriguing given the success of JUUL, a company that spent more resources on social media platforms compared to other tobacco companies,^46^ buttressing that many of JUUL’s earlier marketing strategies unwittingly targeted younger individuals.^13^ Also, although the Tobacco-21 law was passed in Maryland in 2019, restricting the sales of tobacco products, including ENDS, to individuals over the age of 21 years,^47^ almost half of the current users who reported buying their pods in-person were below the age of 21. This is particularly concerning and necessitates stricter enforcement of the Tobacco-21 legislation.

Interestingly, majority of participants reported having seen anti-tobacco ads, although very few engaged with them. Nonetheless, unlike other studies, a vast majority of the participants in our study were well-informed about the nicotine content and the potential harms associated with pod-mod use,^37,48^ with most citing health concerns as their reason for attempting to quit. Majority of the participants in our study had tried to quit, however, considering the high nicotine content of many pod-mods and the “cold turkey” approach to quitting adopted by many participants, it is not surprising that their quit attempts had been unsuccessful. Our findings suggest that quitting ENDS use may be challenging, and the use of nicotine replacement therapy and other prescription medications to this end should be explored. Furthermore, the use of multiple tobacco products observed among current users is in keeping with prior studies and may also be responsible for difficulty quitting.^23,24,30,38^ Finally, only a handful of participants reported ever reading the warning labels on pod-mod packs. While the FDA’s guidelines for including health information on ENDS packaging is important,^49^ a shift towards using graphical health warnings might be more likely to capture the attention of users.

Our findings provide important information on the factors associated with the use of pod-based e-cigarettes among college-aged adults. We provide additional insight into potential avenues for impactful youth education concerning ENDS use, for example, our data demonstrate that college pod-mod users are well-aware of the potential health consequences of these products, therefore, ongoing advertising campaigns on the dangers of pod-mods are unlikely to be effective—different approaches are needed to impact this important population. Additionally, our findings highlight the need for more robust evidence-based cessation support designed for college students trying to quit ENDS use for example, occupational health services on college campuses could be expanded to include ENDS cessation counseling. The need for tighter enforcement of established tobacco laws is also brought to the fore, and additional insight into the impact of current policies and the perception of public health messages will prove invaluable in building further strategies to reduce tobacco use in this population.

A major strength of our study was its careful inclusion of only university students who reported using pod-mods, with multiple safeguards to ensure the integrity of this population. Additionally, few participants were “dual users” of pod-mods and combustible cigarettes, enabling us to focus on a group that were largely exclusive users of pod-mods. Our study however has several limitations. The sample size for our study was modest, and the selection of participants from a college limits the generalizability of our results to other young adults in the population or from other regions or countries. The study’s modest sample size also precludes our determining variation by substantive demographic factors, such as race/ethnicity or sex. Also, the self-reported format of the survey might result in recall bias. The study was observational and cross-sectional, and hence we could not establish the temporality of the associations or establish causal relations.

In conclusion, the use of pod-mods in our study of college-aged young adults was associated with experimentation with multiple tobacco products and addictive behaviors. Social influence played a major role in the uptake and continued use of pod-mods among users, and while many had attempted to quit due to the potential health effects associated with their use, many of the quit attempts were unsuccessful. Our results highlight the need for additional public health strategies and vaping cessation support targeted at college students in the US.

## Data Availability

All data produced in the present study are available upon reasonable request to the authors.

## Declarations

### Ethics approval and consent to participate

This study was approved by the Johns Hopkins School of Medicine Institutional Review Board under IRB number IRB00226738. All participants provided informed consent electronically before proceeding to take the survey. All methods were performed in accordance with the Declaration of Helsinki.

### Availability of data and materials

To protect the privacy of participants, the dataset generated and analyzed during the current study are not publicly available. The fully deidentified dataset is however available from the corresponding author upon reasonable request and with the permission of Johns Hopkins University.

## Acknowledgements

Manuscript has been submitted as pre-print to MedRxiv.

## Authors’ contributions

OHO was involved in study conceptualization and design, data management and analysis, manuscript drafting and editing, and obtaining funding.

MJB was involved in study conceptualization and design, critical review of the manuscript, and obtaining funding.

SMIU was involved in study conceptualization and design, recruitment and consenting of participants and manuscript review.

ADO, OOO, MM were involved in study conceptualization and design and critical review of the manuscript.

EB, OD, AS, APD, OE, EJB were involved in critical review of the manuscript.

