## Supplemental Tables for "Pod-based e-cigarette use among college-aged adults in the United States: A survey on the perception of health effects, sociodemographic correlates, and interplay with other tobacco products"

**SUPPLEMENT TABLES**

Supplement Table 1: Exposure to marketing

Supplement Table 2: Exposure to public health messaging

Supplement Table 3: Perception of harm

Supplement Table 4: Characteristics of PM users by autonomy status per hooked on nicotine checklist

Supplement Table 5: Association between autonomy status and characteristics of PM users

Supplement 6: Questionnaire

**Supplement Table 1: Exposure to marketing**

|  | N (%) |
| --- | --- |
|  | Total, (N=112) |
| When you are using the Internet, how often do you see any ads or promotions for pod-mods? |  |
| - I do not use the internet | 1(0.9) |
| - Never | 48(42.9) |
| - Rarely | 43(38.4) |
| - Sometimes | 19(17.0) |
| - Most of the time | 1(0.9) |
| Pod-mods discussed on social networking account | 44 (39.3) |
| Received information from pod-mod company over past 30 days through... (choose all that apply) |  |
| - E-mail | 9(8.0) |
| - Social network account | 2(1.8) |
| - Some other way | 1(0.9) |
| Heard about sport or sporting event sponsored by or connected with pod-mods in past 6 months | 5(4.5) |
| Heard about music, theater, art, or fashion events sponsored by or connected with pod-mods | 7(6.3) |

**Supplement Table 2: Exposure to public health messaging**

|  | N (%) |
| --- | --- |
|  | Total, (N=112) |
| Recently seen an anti-smoking or anti-tobacco ad on TV or social media | 83(74.1) |
| Discussed content of ad with anyone | 16(14.3) |
| Content discussed |  |
| - The ads were good | 8(7.14) |
| - The ads were not good | 8(7.14) |
| - I should not use e-cigarettes | 10(8.9) |
| - The person I was talking to, or someone else I know should not use e-cigarettes | 6(5.4) |
| Ads are worth remembering |  |
| - Agree | 45(40.2) |
| - Neither agree nor disagree | 20(17.9) |
| - Disagree | 19(17.0) |
| Ads are informative |  |
| - Agree | 49(43.7) |
| - Neither agree nor disagree | 21(18.8) |
| - Disagree | 16(14.3) |
| Ads are convincing |  |
| - Agree | 36(32.2) |
| - Neither agree nor disagree | 26(23.2) |
| - Disagree | 24(21.4) |

**Supplement Table 3: Perception of harm**

|  | N (%) |
| --- | --- |
|  | Total, (N=112) |
| Pod-mod e-cigarettes contain nicotine | 107(95.5) |
| Compared to cigarettes pod-mod e-cigarettes are |  |
| - more harmful | 6(5.4) |
| - equally harmful | 41(36.6) |
| - less harmful | 65(58.0) |
| Compared to cigarettes pod-mod e-cigarettes are |  |
| - more addictive | 44(39.3) |
| - equally addictive | 53(47.3) |
| - less addictive | 15(13.4) |
| What in your view are the main harms, if any, of pod-mod use? (Choose all that apply) |  |
| - There are no harms | 1(0.9) |
| - They may be addictive | 97(86.6) |
| - They may explode or catch fire | 21(18.8) |
| - They may cause breathing/respiratory problems | 94(83.9) |
| - They may cause cancer | 70(62.5) |
| - The chemicals in the liquid may be harmful | 85(75.9) |
| - The nicotine in the liquid may be harmful | 57(50.9) |
| - They reinforce the smoking habit | 72(64.3) |
| - They may cause heart disease | 39(34.8) |
| - There is not enough quality control/regulation | 60(53.6) |
| There has not been enough research done to understand all the possible harms | 83(74.1) |

**Supplement Table 4: Characteristics of PM users by autonomy status per hooked on nicotine checklist**

|  | N (%) | | | |
| --- | --- | --- | --- | --- |
|  | Total  (N=112) | Full autonomy  (N=38) | Reduced autonomy^a^  (N=62) | p-value |
| PM use category |  |  |  |  |
| - Current user | 45(40.2) | 7(18.4) | 38(61.3) | 0.00 |
| - Non-current user | 67(59.8) | 31(81.6) | 24(38.7) |  |
| Most frequently used brand of pod e-cigarettes^b^ |  |  |  |  |
| - JUUL | 51(45.5) | 12(31.6) | 39(62.9) | 0.00 |
| - Suorin | 3(2.7) | 2(5.3) | 1(1.6) |  |
| - Joytech | 1(0.9) | 0(0) | 1(1.6) |  |
| - Puff Bar | 3(2.7) | 1(2.6) | 2(3.2) |  |
| - Vuse | 1(0.9) | 0(0) | 1(1.6) |  |
| - Other | 2(1.8) | 0(0) | 2(3.2) |  |
| - None | 51(45.5) | 23(60.5) | 16(25.8) |  |
| Pod flavor used most frequently^b^ |  |  |  |  |
| - Virginia tobacco | 2(1.8) | 0(0) | 2(3.2) | 0.02 |
| - Mint | 10(8.9) | 4(10.5) | 6(9.7) |  |
| - Mango | 5(4.5) | 2(5.3) | 3(4.8) |  |
| - Classic tobacco | 1(0.9) | 0(0) | 1(1.6) |  |
| - Menthol | 20(17.9) | 2(5.3) | 18(29.0) |  |
| - Fruit | 10(8.9) | 3(7.8) | 7(11.3) |  |
| - None | 64(57.1) | 27(71.1) | 25(40.3) |  |
| Other nicotine product ever use (select all) |  |  |  |  |
| - Cigarettes | 71(63.4) | 19(50) | 48(77.4) | 0.00 |
| - Cigar | 44(39.3) | 13(34.2) | 28(45.2) | 0.311 |
| - Cigarillo | 34(30.4) | 7(18.4) | 25(40.3) | 0.04 |
| - Blunt | 67(59.8) | 19(50) | 44(70.9) | 0.02 |
| - Pipe | 20(17.9) | 6(15.8) | 14(22.6) | 0.16 |
| - Hookah | 43(38.4) | 10(26.3) | 32(51.6) | 0.00 |
| - Snus pouches | 6(5.4) | 3(7.9) | 3(4.8) | 0.55 |
| - Loose snus/moist snuff | 6(5.4) | 1(2.6) | 5(8.1) | 0.34 |

^a^ defined as HONC score>=1

^b^ In the past 30 days

**Supplement Table 5: Association between autonomy status and characteristics of PM users**

|  | Reduced autonomy^a^ | |
| --- | --- | --- |
|  | Unadjusted  OR(95%CI) | Adjusted^b^  OR(95%CI) |
| PM use category |  |  |
| - Current user | 4.67(1.83,11.95) | 4.52(1.76,11.64) |
| - Non-current user | ref | ref |
| Most frequently used brand of pod e-cigarettes^c^ |  |  |
| - JUUL | 2.67(1.14,6.25) | 2.56(1.08,6.03) |
| - Suorin | 0.41 (0.03,4.82) | 0.31(0.02,3.94) |
| - Joytech | -- | -- |
| - Puff Bar | 1.64(0.13,9.29) | 1.68(0.14,19.98) |
| - Vuse | -- | -- |
| - Other | -- | -- |
| - None | ref | ref |
| Pod flavor used most frequently^c^ |  |  |
| - Virginia tobacco | -- | -- |
| - Mint | 1.09(0.28,4.26) | 1.09(0.27,4.42) |
| - Mango | 1.09(0.17,7.01) | 1.01(0.15,6.64) |
| - Classic tobacco | -- | -- |
| - Menthol | 6.57(1.40,30.72) | 6.52(1.38,30.89) |
| - Fruit | 1.70(0.40,7.19) | 1.49(0.34,6.58) |
| - None | ref | ref |

^a^ defined as HONC score>=1

^b^adjusted for age and sex

**Supplement 6: Questionnaire**

Do you agree to participate in this survey? Y/N

Please enter unique ID code: _____________

DEMOGRAPHIC INFORMATION

1) What is your year of birth? (yyyy) _________
   a. [ ] Don’t know
2) About how old are you? ___________
   a. [ ] Don’t know
3) What was your biological sex assigned at birth?
   a. [ ] Female
   b. [ ] Male
   c. [ ] Intersex
   d. [ ] None of these describe me
   e. [ ] Prefer not to answer
4) What is the highest level of school you have completed?
   a. [ ] None
   b. [ ] Middle to High School (specify grade) ____________
   c. [ ] High School Graduate
   d. [ ] Some college, no degree
   e. [ ] Bachelor’s degree (e.g. BA, AB, BS, BBA)
   f. [ ] Master’s degree (e.g. MA,MS, MEng, MEd, MBA)
   g. [ ] Professional school degree (e.g. MD,DDS, DVM, JD)
5) What race or races do you consider yourself to be?
   a. [ ] American Indian or Alaska native
   b. [ ] Asian
   c. [ ] Black or African-American
   d. [ ] Native Hawaiian or Pacific Islander
   e. [ ] White
   f. [ ] Other
6) What ethnicity do you consider yourself to be?
   a. [ ] Hispanic
   b. [ ] Non-Hispanic
7) In what category is your best estimate of the total family income of all family members from all sources, before taxes, in last calendar year?
   a. [ ] Less than $35,000
   b. [ ] Above $35,000 but less than $50,000
   c. [ ] Above $50,000 but less than $75,000
   d. [ ] Above $75,000 but less than $100,000
   e. [ ] $100,000 or more
   f. [ ] Don’t know

**E-CIGARETTE USE**

The next questions are about electronic cigarettes, often called e-cigarettes. E-cigarettes look like regular cigarettes, but are battery-powered and produce vapor instead of smoke. They are also referred to as “E-cigs” “e-hookahs” “mods” “vape pens” “vapes” “tank systems” and “electronic nicotine delivery systems”. Using an e-cigarette is sometimes called “vaping” or “JUULing”. Some common brands include NJOY®, Blu™, JUUL, Vuse, Mark-Ten, Logic and eGO^26^and Smoking Everywhere.

Picture: <https://upload.wikimedia.org/wikipedia/commons/d/dc/CDC_electronic_cigarettes_October_2015_%28cropped%29.png>

1) Which of the following electronic nicotine products have you used? Choose all that apply.
   a. [ ]E-cigarette including vape pens, hookah pens, personal vaporizers, and mods
   b. [ ]E-cigar
   c. [ ]E-pipe
   d. [ ]E-hookah
   e. [ ]Something else (specify) ____________________
2) Do you now use e-cigarettes…
   a. [ ] Everyday
   b. [ ] Some days
   c. [ ] Not at all
3) In the past 30 days, have you used an e-cigarette, even one or two times?
   a. [ ] Yes
   b. [ ] No
4) Have you completely quit using e-cigarettes?
   a. [ ] Yes
   b. [ ] No

**POD-BASED E-CIGARETTE USE**

The next questions are about pod-based e-cigarettes, called **pod-mods**. Brands include JUUL, MyBlu, SMPO, MLV, Joyetech, Suorin and Aspire. Pod-Mods use replaceable cartridges containing e-liquid called “pods”.

1) Do you now use pod-mods …
   a. [ ] Everyday
   b. [ ] Some days
   c. [ ] Not at all
2) What was the first pod flavor you ever used?^26^
   a. [ ] Virginia Tobacco
   b. [ ] Mint
   c. [ ] Mango
   d. [ ] Crème
   e. [ ] Cucumber
   f. [ ] Classic tobacco
   g. [ ] Menthol
   h. [ ] Fruit
   i. [ ] other……Please specify ___________
3) In the past 30 days, have you used pod-mods, even one or two times?
   a. [ ] Yes
   b. [ ] No
4) During the past 30 days which flavor of refill pod have you used the most often? (select one)^26^
   a. [ ] Virginia Tobacco
   b. [ ] Mint
   c. [ ] Mango
   d. [ ] Crème
   e. [ ] Cucumber
   f. [ ] Classic tobacco
   g. [ ] Menthol
   h. [ ] Fruit
   i. [ ] None
5) What brand of pod-mods have you used most frequently in the last 30 days?
   a. [ ] JUUL
   b. [ ] Suorin
   c. [ ] Blu
   d. [ ] Ruby
   e. [ ] SMPO
   f. [ ] MLV
   g. [ ] Joytech
   h. [ ] Aspire
   i. [ ] Mistic
   j. [ ] None
   k. [ ] Other: __________________
6) How many pods do/did you typically finish in 30 days?^22^
   a. [ ] Less than 1 pod
   b. [ ] 1 to 2 pods (about ½ pack)
   c. [ ] 3 to 4 pods (about ½ to 1 pack)
   d. [ ] 5 to 12 pods (more than 1 pack but less than 3 packs)
   e. [ ] 13 to 19 pods (more than 2½ packs but less than 5 packs)
   f. [ ] 20 or more (5 packs or more)
7) How old were you when you first tried a pod-mod? ________years
8) How old were you when you first started using pod-mods fairly regularly? ________years
9) Do you own a pod-mod?
   a. [ ] Yes
   b. [ ] No
10) About how much did you pay for your pod-mod? Do not include the cost of additional cartridges or accessories unless they were included in a starter kit.
    a. [ ] Less than $10
    b. [ ] $10 to $30
    c. [ ] $31 to $40
    d. [ ] $41 to $50
    e. [ ] More than $50
    f. [ ] Not applicable
11) How do/did you usually buy pods for yourself?
    a. [ ] In person
    b. [ ] From the internet
    c. [ ] By telephone
    d. [ ] I do/did not buy my own pods
12) Why did you start using pod-mods?^24^ (choose all that apply)
    a. [ ] I was curious
    b. [ ] Friends/Family use or gave me one to try
    c. [ ] They are less harmful to me compared to smoking regular cigarettes
    d. [ ] They are less harmful to others
    e. [ ] They do not smell bad
    f. [ ] They do not bother others
    g. [ ] They are affordable
    h. [ ] They come in flavors I like
    i. [ ] I like the ads for them
    j. [ ] They helped me quit/reduce smoking
    k. [ ] I can use them where I can’t smoke
    l. [ ] Using them feels like smoking
    m. [ ] Using them helps with cravings
13) Where did you first try a pod-mod?
    a. [ ] At home or a friend’s home
    b. [ ] In school
    c. [ ] At a party, night-club or concert
    d. [ ] At a bar or hookah lounge
    e. [ ] At a community event (e.g. a food fair, baseball game, carnival)
    f. [ ] Other: please specify______________
14) How soon after you wake up do/did you take your first puff of a pod-mod?
    a. [ ] Within 5 minutes
    b. [ ] 6-30 minutes
    c. [ ] 31-60 minutes
    d. [ ] After 60 minutes
15) Do/did you find it difficult to refrain from using it in places where it is forbidden e.g. school, church?
    a. [ ] Yes
    b. [ ] No
16) Do/did you smoke more frequently during the first hours after waking than during the rest of the day?
    a. [ ] Yes
    b. [ ] No
17) Have you ever felt like you were addicted to using pod-mods?^23^
    a. [ ] Yes
    b. [ ] No
18) Do/did you ever have strong cravings for pod-mods?^23^
    a. [ ] Yes
    b. [ ] No
19) Are you considering quitting pod-mod use during the next 6 months?
    a. [ ] Yes
    b. [ ] No
20) In the past, have you ever made a serious attempt to quit pod-mods? That is, have you stopped using them for at least one day or longer because you were trying to quit?
    a. [ ] Yes
    b. [ ] No
21) What was the longest length of time you stopped using pod-mods because you were trying to quit?
    a. [ ] Less than 1 week
    b. [ ] 1 to 3 weeks
    c. [ ] 1 to 2 months
    d. [ ] 3 to 11 months
    e. [ ] 1 to 4 years
    f. [ ] Not applicable
22) How old were you when you most recently quit using pod-mods? ________ years
23) Why did you decide to stop using pod-mods?
    a. [ ] I was just experimenting
    b. [ ] I did not feel like using them
    c. [ ] I did not like the taste
    d. [ ] It cost too much
    e. [ ] It didn’t help me quit or cut back smoking
    f. [ ] It didn’t help with my cravings
    g. [ ] I was concerned about the health risks
    h. [ ] The quality was poor
    i. [ ] I did not like the side effects
    j. [ ] I saw ads on potential dangers of vaping
    k. [ ] Not applicable
24) Thinking of the most recent time you quit using pod-mods, did you use any of the following products to help you quit?
    a. [ ] Nicotine gum
    b. [ ] Nicotine patch
    c. [ ] Nicotine nasal spray, inhaler, lozenge or tablet
    d. [ ] Prescription pill such as Zyban®, Bupropion, or Wellbutrin® (Zyban® is a registered trademark, GlaxoSmithKline; Wellbutrin® is a registered trademark, GlaxoSmithKline)
    e. [ ] None of these
    f. [ ] Not applicable
25) If you have tried to quit pod-mod use/ have not used it for a while: Did you find it hard to concentrate because you could not use a pod-mod?^23^
    a. [ ] Yes
    b. [ ] No
26) If you have tried to quit pod-mod use/ have not used it for a while: Did you feel a strong need/urge to use pod-mods?^23^
    a. [ ] Yes
    b. [ ] No
27) If you have tried to quit pod-mod use/ have not used it for a while: Did you feel nervous, anxious or irritable because you could not use a pod-mod?^23^
    a. [ ] Yes
    b. [ ] No
28) During the past 12 months, did any doctor, dentist, nurse, or any other health professional advice you to quit using pod-mods?
    a. [ ] Yes
    b. [ ] No, was not advised to quit
    c. [ ] No, did not see a health professional in past 12 months
    d. [ ] No, did not smoke in past 12 months
29) In the past 30 days, how often, if at all, have you read or looked closely at the health warnings on pod-mod packages?
    a. [ ] Never
    b. [ ] Rarely
    c. [ ] Sometimes
    d. [ ] Often
    e. [ ] Very often
30) Compared to cigarettes, do you think pod-mod e-cigarette is more harmful, less harmful or equally harmful?^25^
    a. [ ] More harmful than tobacco cigarettes
    b. [ ] Equally harmful
    c. [ ] Less harmful
31) Do you think pod-mod e-cigarettes contain nicotine?
    a. [ ] Yes
    b. [ ] No
32) Compared to cigarettes, do you think pod-mod e-cigarette is more addictive, less addictive or equally addictive?^25^
    a. [ ] More addictive
    b. [ ] Equally addictive
    c. [ ] Less addictive
33) What in your view are the main harms, if any, of pod-mod use?^25^ (select all that apply)
    a. [ ] There are no harms
    b. [ ] They may be addictive
    c. [ ] They may explode or catch fire
    d. [ ] They may cause breathing/respiratory problems
    e. [ ] They may cause cancer
    f. [ ] The chemicals in the liquid may be harmful
    g. [ ] The nicotine in the liquid may be harmful
    h. [ ] They reinforce the smoking habit
    i. [ ] They may cause heart disease
    j. [ ] There is not enough quality control/regulation
    k. [ ] There has not been enough research done to understand all the possible harms
34) Does anyone who lives with you use a pod-mod?
    a. [ ] Yes
    b. [ ] No
35) Of the five closest friends or acquaintances that you spend time with on a regular basis, how many of them use a pod-mod? _______________
36) Thinking about the people who are important to you, how would you describe their opinion on using pod-mods?
    a. [ ] Very positive
    b. [ ] Positive
    c. [ ] Neither positive nor negative
    d. [ ] Negative
    e. [ ] Very negative
37) How would you describe most people’s opinion of using pod-mods?
    a. [ ] Very positive
    b. [ ] Positive
    c. [ ] Neither positive nor negative
    d. [ ] Negative
    e. [ ] Very negative

**Pod-mod Marketing**

1) When you are using the Internet, how often do you see any ads or promotions for pod-mods?
   a. [ ] I do not use the internet
   b. [ ] Never
   c. [ ] Rarely
   d. [ ] Sometimes
   e. [ ] Most of the time
   f. [ ] Always
2) Has anyone discussed pod-mods on your Facebook, Google Plus, Twitter, or other social networking account?
   a. [ ] Yes
   b. [ ] No
3) During the past 30 days, did any pod-mod company send you information (other than coupons) through… (choose all that apply)
   a. [ ] The mail
   b. [ ] E-mail
   c. [ ] Social Networks (e.g. Facebook, Twitter)
   d. [ ] Text message
   e. [ ] Some other way
   f. [ ] I did not receive any information from a tobacco company
4) Thinking about the last 6 months have you seen or heard about any sport or sporting event that is sponsored by or connected with pod-mods?
   a. [ ] Yes
   b. [ ] No
5) Have you heard about any music, theater, art, or fashion events that are sponsored by or connected with pod-mods?
   a. [ ] Yes
   b. [ ] No

**PUBLIC HEALTH MESSAGES**

1) Have you recently seen an anti-smoking or anti-tobacco ad on TV or social media? (either brief verbal, text, screenshot or video)
   a. [ ] Yes
   b. [ ] No
2) Did you talk to anyone about this ad?
   a. [ ] Yes
   b. [ ] No
3) When you talked about this ad, did you talk about any of the following topics? (choose all that apply)
   a. [ ] The ads were good
   b. [ ] The ads were not good
   c. [ ] I should not use e-cigarettes
   d. [ ] The person I was talking to, or someone else I know should not use e-cigarettes

How much do you agree or disagree with each the following statements?

4) These ads are worth remembering
   a. [ ] Strongly disagree
   b. [ ] Disagree
   c. [ ] Neither agree nor disagree
   d. [ ] Agree
   e. [ ] Strongly agree
5) These ads are informative
   a. [ ] Strongly disagree
   b. [ ] Disagree
   c. [ ] Neither agree nor disagree
   d. [ ] Agree
   e. [ ] Strongly agree
6) These ads are convincing
   a. [ ] Strongly disagree
   b. [ ] Disagree
   c. [ ] Neither agree nor disagree
   d. [ ] Agree
   e. [ ] Strongly agree

**USE OF OTHER TOBACCO PRODUCTS**

1) Have you ever smoked a cigarette, even one or two puffs?
   c. [ ] Yes
   d. [ ] No (go to question 4)
2) Do you now smoke cigarettes…
   a. [ ] Everyday
   b. [ ] Some days
   c. [ ] Not at all

The next questions are about traditional cigars, cigarillos, and filtered cigars. These products go by lots of different names, so please use these descriptions and photos to understand what they are. Traditional cigars contain tightly rolled tobacco that is wrapped in a tobacco leaf. Some common brands of cigars include Macanudo®, Romeo y Julieta®, and Arturo Fuente®, but there are many others.

Cigarillos and filtered cigars are smaller than traditional cigars. They are usually brown. Some are the same size as cigarettes, and some come with plastic or wood tips. Some common brands are Black & Mild®, Swisher Sweets®, Dutch Masters®, Phillies Blunts®, Prime Time®, and Winchester®.

3) Have you ever smoked a traditional cigar, even one or two puffs?
   a. [ ] Yes
   b. [ ] No
4) Have you ever smoked a cigarillo or filtered cigar, even one or two puffs?
   a. [ ] Yes
   b. [ ] No
5) Sometimes people take tobacco out of a cigar, cigarillo or filtered cigar and replace it with marijuana. This is sometimes called a "blunt." Have you ever smoked part or all of a cigar, cigarillo or filtered cigar with marijuana in it?
   a. [ ] Yes
   b. [ ] No
6) Do you now smoke [Cigars/Cigarillos / Blunts]…
   a. [ ] Everyday
   b. [ ] Some days
   c. [ ] Not at all
7) Have you ever smoked a pipe filled with tobacco, even one or two puffs?
   a. [ ] Yes
   b. [ ] No
8) Do you now smoke a pipe filled with tobacco…
   a. [ ] Everyday
   b. [ ] Some days
   c. [ ] Not at all
9) Have you ever smoked tobacco in a hookah, even one or two puffs?
   a. [ ] Yes
   b. [ ] No
10) Do you now smoke hookah…
    a. [ ] Everyday
    b. [ ] Some days
    c. [ ] Not at all
11) Have you ever used any of the following smokeless tobacco products, even one or two times? Choose all that apply.
    a. [ ] Snus pouches
    b. [ ] Loose snus, moist snuff, dip, spit, or chewing tobacco
    c. [ ] I have never used a smokeless tobacco product
12) Do you now use any of the above listed smokeless tobacco products….
    a. [ ] Everyday
    b. [ ] Some days
    c. [ ] Not at all
13) Have you ever used dissolvable tobacco products, such as Ariva®, Stonewall™, or Camel Orbs, Sticks, or Strips, even one or two times?
    a. [ ] Yes
    b. [ ] No
14) Do you now use dissolvable tobacco…
    a. [ ] Everyday
    b. [ ] Some days
    c. [ ] Not at all
